# Sixteen Days Undetected: Growth Dynamics and the Case for Pre-Positioned Response Capacity in the 2026 Bundibugyo Virus Disease Outbreak, Democratic Republic of the Congo

**DOI:** 10.64898/2026.08.12.26360240

**Authors:** Johan G.L. Verheyden, Celestin Nzanzu Mudogo

**Author notes:** Corresponding author: Celestin Nzanzu Mudogo —, Johan G.L. Verheyden —.

## Abstract

**Objectives:** To estimate early growth rate, back-calculate transmission onset, and characterise the case-fatality trajectory of the 2026 Bundibugyo virus disease (BDBV) outbreak in the Democratic Republic of the Congo, the largest recorded BDBV outbreak to date.

**Design or methods:** We analysed a corrected daily surveillance series (65 observations, 14 May–27 July 2026) using non-linear least-squares regression and a Bayesian Poisson growth model fitted by Markov chain Monte Carlo, with five sensitivity analyses.

**Results:** Early confirmed cases grew at 0·1261 per day (95% CI 0·0885–0·1636), a doubling time of 5·50 days (4·24–7·83), three-fold faster than previous BDBV outbreaks (15–18 days). Bayesian back-calculation placed transmission onset on 19 April 2026 (95% highest-density interval 9–27 April), 16 days before the WHO alert and 25 days before laboratory confirmation. Confirmed case-fatality ratio rose from 12·1% to 44·3%; a higher ratio among suspected than confirmed cases on 21 May (23·6% vs 10·8%; p=0·0080) supported progressive reclassification rather than increasing virulence.

**Conclusions:** Rapid BDBV growth leaves little time for outbreak-triggered mobilisation. Sentinel alerts based on unexplained healthcare-worker death clusters, together with pre-positioned surveillance, diagnostic, and response capacity, could reduce avoidable amplification before confirmation.

## Introduction

Bundibugyo virus (BDBV; Orthoebolavirus bundibugyoense) was first identified in 2007 during an outbreak of haemorrhagic fever in Bundibugyo District, western Uganda.^1^ Two outbreaks preceded 2026: the index Uganda epidemic (2007–2008; 116–149 cases; CFR 25–34%)^2,3^ and a subsequent epidemic in Isiro, Democratic Republic of the Congo (DRC; 2012; 52–62 cases; CFR 49–54%).^4,5^ Both were relatively contained, with doubling times of 15–18 days. Neither triggered the kind of pre-positioned response infrastructure that now exists for Zaire ebolavirus (EBOV), for which ring vaccination has demonstrably reduced transmission in this same region.^6^ For BDBV, no licensed vaccine or therapeutic exists; response depends entirely on early detection and non-pharmaceutical intervention.

The 2026 outbreak, declared 15 May 2026 by the DRC Ministry of Public Health, is a categorical departure from this history. Centred initially in Ituri Province — a region with active armed conflict, mining-related displacement, and prior EBOV transmission during the 2018–2020 North Kivu and Ituri epidemic — it expanded across three DRC provinces and generated secondary transmission in Uganda.^7^ Phylogenetic analysis confirmed the 2026 virus as genetically distinct from the 2007 and 2012 lineages, establishing an independent spillover event.^8^ This is the first documented BDBV epidemic to sustain transmission in a major urban centre: Bunia, capital of Ituri Province, population approximately 900,000.

For health systems planning around filovirus species that lack dedicated countermeasures, three questions bear directly on operational readiness. First, the outbreak reached 1,000 confirmed cases within approximately 33 days of the first laboratory confirmation9 — compared with 245 days for the 2018–2020 EBOV epidemic in the same region.6 Is this outbreak intrinsically faster, or did a longer undetected transmission period precede recognition? Second, the first WHO Disease Outbreak Notification (16 May 2026) reported 246 suspected cases and 80 suspected deaths against only eight confirmed cases,10 implying substantial pre-declaration accumulation. Third, the case fatality ratio (CFR) rose from 12·1% to 44·3% over the observation period — a trajectory that could reflect either worsening severity or an ascertainment artefact, with materially different implications for clinical resource planning.

Growth-rate estimation answers the first two questions directly: doubling time determines how fast response capacity must scale,^11,12^ and back-calculating outbreak onset from observed case counts is an established approach^13^ for quantifying pre-recognition transmission, documented in multiple prior filovirus epidemics.^14,15^ Decomposing the CFR trajectory into genuine severity change versus ascertainment artefact is essential for both risk communication and clinical triage planning.^16,17^

This analysis addresses three questions with direct operational relevance: how much faster is the 2026 outbreak growing than prior BDBV experience; how long did transmission go undetected before international alert, and what would earlier detection have been worth in cases averted; and what explains the observed rise in CFR. We apply both classical regression and Bayesian hierarchical inference, using their concordance as an internal validity check, and translate the results directly into implications for surveillance-protocol design and pre-positioned response capacity.

## Methods

### Data sources

Cumulative confirmed case counts and deaths were extracted from a corrected daily surveillance series (65 observations, 14 May–27 July 2026) drawn from the INRB-UMIE BDBV2026-Data repository of digitised DRC INSP situation reports.18 This corrected series superseded an earlier extraction from WHO AFRO Weekly External Situation Reports, which we found on reanalysis to contain date-alignment and transcription errors; all results reported here use the corrected daily series throughout, independently verified against the underlying INSP SitRep extraction. The series was extended through 27 July by aggregating the same repository’s health-zone-level records to national daily totals; every resulting checkpoint reproduced independently published figures exactly (15 July: 2,124 cases/828 deaths, WHO Disease Outbreak News; 18 July: 2,344/930, NICD; 27 July: 3,360/1,487, ECDC). Ten reporting dates (15–16 May, 12 June, 26 June, 28 June, 12 July, 14 July, 16 July, 21 July, and 24 July 2026) lack confirmed-case counts in the source and were excluded rather than interpolated. Epidemiological context and the 21 May 2026 cross-classification data were taken from Mwamba et al. and Tonen-Wolyec and Bélec.7,19 All data derive from published surveillance reports and peer-reviewed literature; ethics approval was not required.

### Growth models

Cumulative case counts C(t) were modelled as C(t) = C₀·exp(r·t), where r is the exponential growth rate and doubling time Tᵈ = ln(2)/r. The early-phase model (14–27 May, pre-discontinuity) was fitted by non-linear least squares to 12 daily observations; a logistic model, C(t) = K / [1 + ((K − C₀)/C₀)·exp(−r·t)], was fitted to the full 65-observation series and compared against the exponential model by Akaike Information Criterion.

### Bayesian back-calculation

A Bayesian hierarchical model (Poisson likelihood, weakly informative Normal priors,^20^ fitted by Markov Chain Monte Carlo in PyMC3^21^) estimated growth rate r, initial case count C₀, and t_back — the number of days before the first situation report that transmission began — with full posterior uncertainty propagation. Convergence was assessed by Gelman–Rubin R^ and posterior predictive checks. Full prior specifications, MCMC diagnostics, and five pre-specified sensitivity analyses varying priors, fitting windows, and the observation-model distribution are reported in the appendix, all rerun on the corrected daily series.

### Case fatality ratio

CFR was calculated at each reporting date. At the earliest date with disaggregated confirmed and suspected case and death data (21 May 2026),^7^ we compared confirmed and suspected CFR by two-sample z-test, and examined the temporal CFR trajectory for consistency with progressive reclassification of suspected deaths into the confirmed category — the expected pattern under ascertainment lag — versus an irregular pattern consistent with genuine severity change.

## Results

### Outbreak timeline

On 5 May 2026, WHO received alert of unusual deaths of unknown aetiology in Mongbwalu Health Zone, Ituri Province, including four healthcare workers who died within four days.10 Rapid response teams deployed on 11 May; Bundibugyo virus was confirmed in eight of 13 testable samples at INRB Kinshasa on 14 May. The DRC Ministry of Public Health declared the outbreak on 15 May, the day Uganda confirmed an imported case in Kampala. WHO declared a public health emergency of international concern on 17 May; Africa CDC declared a Public Health Emergency of Continental Security the following day.10,22 By 10 July 2026, 1,873 confirmed cases and 672 confirmed deaths had been reported across 25 health zones in three DRC provinces; by 27 July, 3,360 confirmed cases and 1,487 deaths (CFR 44·3%) had been reported nationally. Supplementary Table S1 (appendix) presents the full timeline.

### Growth rate

The Bayesian posterior growth rate, median r = 0·1334 per day (95% HDI 0·1022–0·1697), is broadly concordant (5·8% relative difference), providing cross-methodological validation. Over the full 65-observation series (14 May–27 July), growth rate fell to r = 0·0364 per day (Tᵈ = 19·02 days), and a logistic model was preferred by AIC (K ≈ 6,166), confirming substantial deceleration as response capacity scaled — though we treat this plateau estimate as provisional: successive one-week extensions of this series have shifted the fitted asymptote from K ≈ 2,333 to K ≈ 3,002 to K ≈ 6,166 (29% then 105% change), exceeding even the sensitivity a companion forecasting analysis found between defensible model specifications on a fixed dataset; see Appendix A6 for the full extension history.Fitted to the 12 pre-discontinuity daily observations (14–27 May), non-linear regression yielded r = 0·1261 per day (95% CI 0·0885–0·1636), a doubling time of 5·50 days (95% CI 4·24–7·83; RZ = 0·867). Table 1 presents full model comparisons; MCMC convergence diagnostics and the five sensitivity analyses are reported in the appendix, where results are broadly but not completely robust, with two of five variants implying a modestly later onset.

**Table 1.** Classical regression model estimates for exponential growth rate, doubling time, and model fit. ··=not applicable to that model. *RZ on log scale, not directly comparable with untransformed models. †Doubling time is not constant for a logistic curve (growth decelerates as cases approach K), so is not reported for this model; K (final size) is reported instead. NLS = non-linear least squares; OLS = ordinary least squares. K estimates should be read with particular caution: this figure has more than doubled across two successive one-week extensions of the data window (2333→3002→6166; Appendix A6) and is reported here for model-comparison completeness, not as a dependable final-size projection.

| Model | Data period | n | $r$ (day<sup>-1</sup>) | $T^d$ (days) | K (final size) | $R^2$ |
| --- | --- | --- | --- | --- | --- | --- |
| Exponential NLS | 14–27 May (early) | 12 | 0·1261<br>(0·0885–0·1636) | 5·50<br>(4·24–7·83) | .. | 0·867 |
| Exponential log-OLS | 14–27 May (early) | 12 | 0·2119 | 3·27 | .. | 0·842* |
| Exponential NLS | 14 May–27 Jul (full) | 65 | 0·0364<br>(0·0347–0·0382) | 19·02<br>(18·16–19·97) | .. | 0·978 |
| Logistic NLS | 14 May–27 Jul (full) | 65 | 0·0498<br>(0·0440–0·0556) | ..† | 6166<br>(4413–7920) | 0·985 |

**Figure 1.**
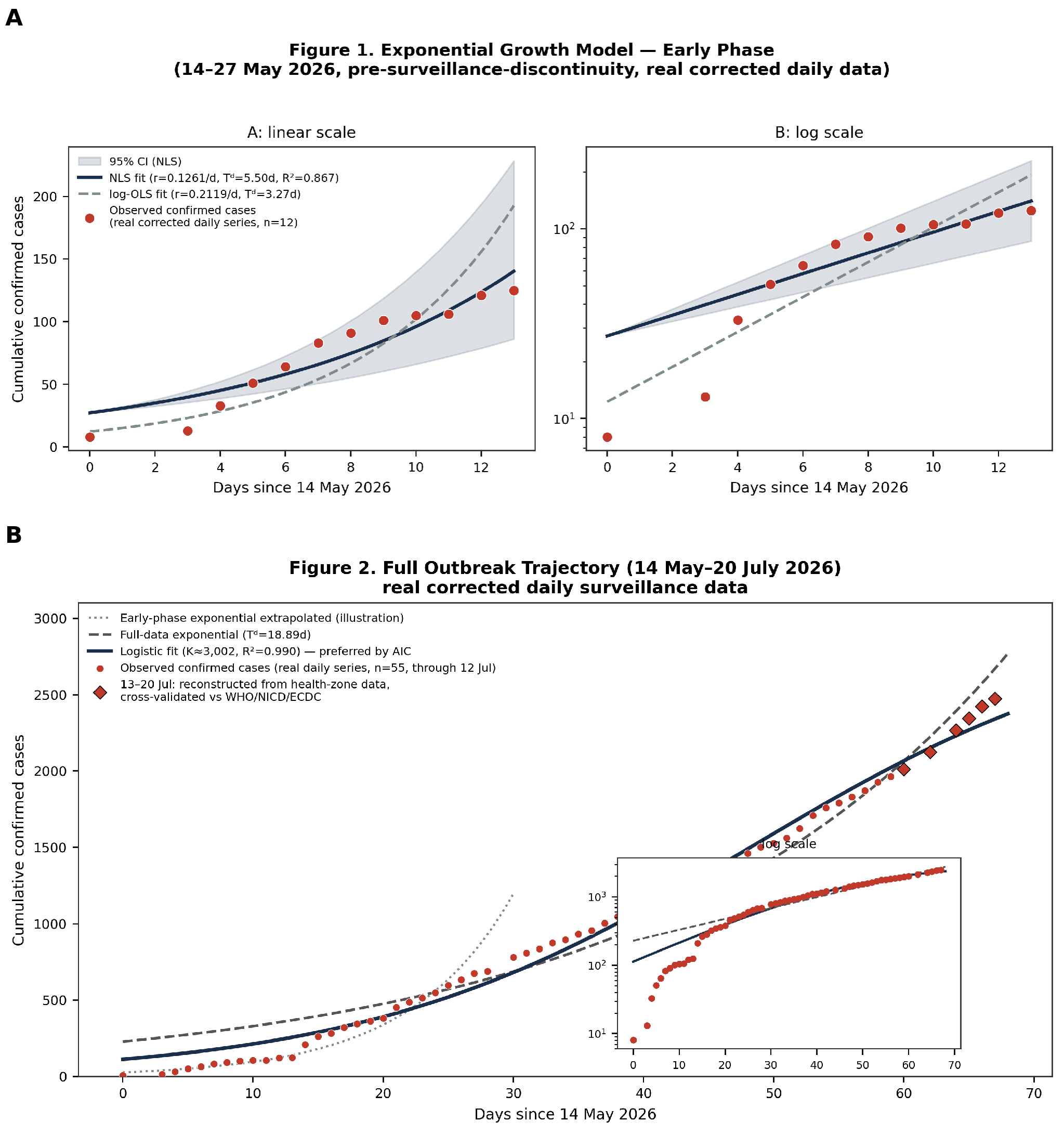
Growth models fitted to real corrected daily surveillance data (rebased, cut-off 27 July 2026). Panel A: exponential growth model, early phase (14–27 May 2026, pre-discontinuity; n=12); NLS fit (r = 0.1261 day⁻¹, Tᵈ = 5.50 days, RZ = 0.867) and log-linear OLS fit, with 95% CI; red circles: observed confirmed cases, independently verified against the underlying INSP daily extraction. Panel B: full outbreak trajectory (14 May–27 July 2026; n=65); early-phase exponential extrapolated (light dashed); full-data exponential (dark dashed; Tᵈ = 19.02 days); logistic fit preferred by AIC (K = 6166; see Appendix A6 — this estimate has more than doubled across two successive one-week cut-off extensions and should be read as illustrative of curve shape, not as a dependable final-size projection); open circles (21–27 July): reconstructed from health-zone-level data, cross-validated against ECDC published figures.

### Back-calculated onset and the cost of detection lag

The Bayesian back-calculation places outbreak onset at 19 April 2026 (95% HDI 9–27 April) — 16 days before the WHO alert (5 May) and 25 days before laboratory confirmation (14 May). The posterior probability that transmission preceded the WHO alert exceeded 99·9%. At a doubling time of 5·5 days, this detection lag alone corresponds to approximately six-fold case growth before international response mechanisms activated: at the WHO alert date, the model implies 4–13 circulating cases (median 8); by declaration on 15 May, 20–38 cases (median 28). Table 2 compares classical and Bayesian estimates of onset, which are closely concordant.

**Table 2.**
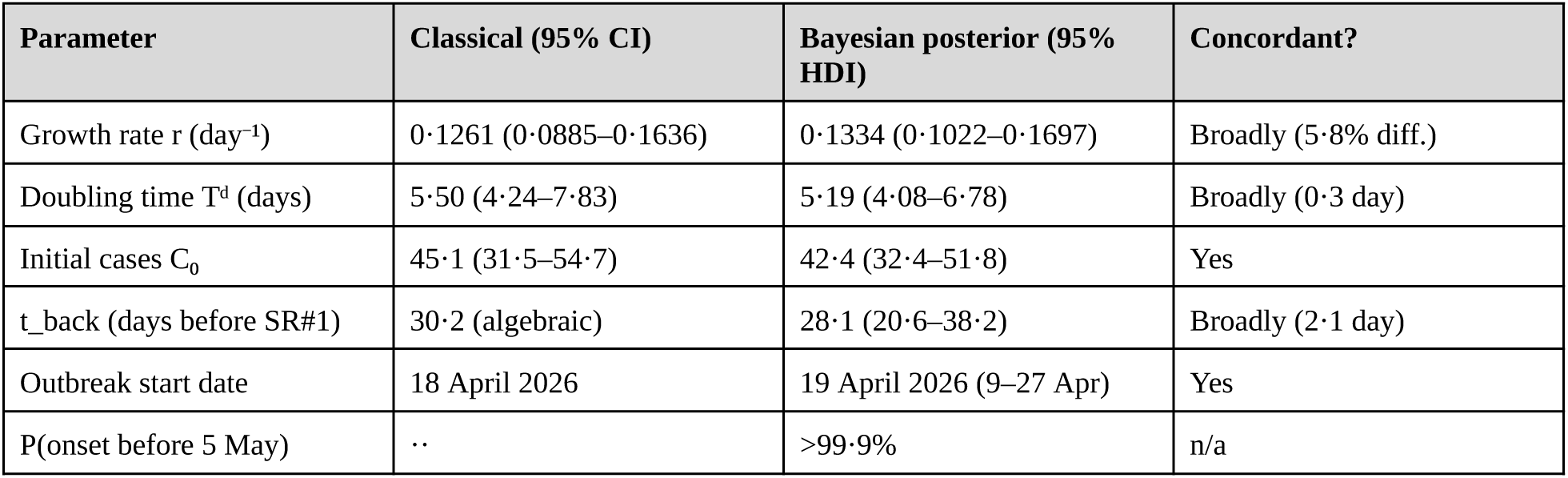
*Comparison of classical regression and Bayesian posterior estimates for growth rate, doubling time, and back-calculated outbreak onset. ··=not applicable (a posterior probability has no classical-estimation analogue). HDI = highest density interval; 16,000 posterior draws (4 chains × 4,000 draws)*.

| Parameter | Classical (95% CI) | Bayesian posterior (95% HDI) | Concordant? |
| --- | --- | --- | --- |
| Growth rate $r$ ($\text{day}^{-1}$) | 0.1261 (0.0885–0.1636) | 0.1334 (0.1022–0.1697) | Broadly (5.8% diff.) |
| Doubling time $T^d$ (days) | 5.50 (4.24–7.83) | 5.19 (4.08–6.78) | Broadly (0.3 day) |
| Initial cases $C_0$ | 45.1 (31.5–54.7) | 42.4 (32.4–51.8) | Yes |
| $t_{\text{back}}$ (days before SR#1) | 30.2 (algebraic) | 28.1 (20.6–38.2) | Broadly (2.1 day) |
| Outbreak start date | 18 April 2026 | 19 April 2026 (9–27 Apr) | Yes |
| P(onset before 5 May) | .. | >99.9% | n/a |

**Figure 2.**
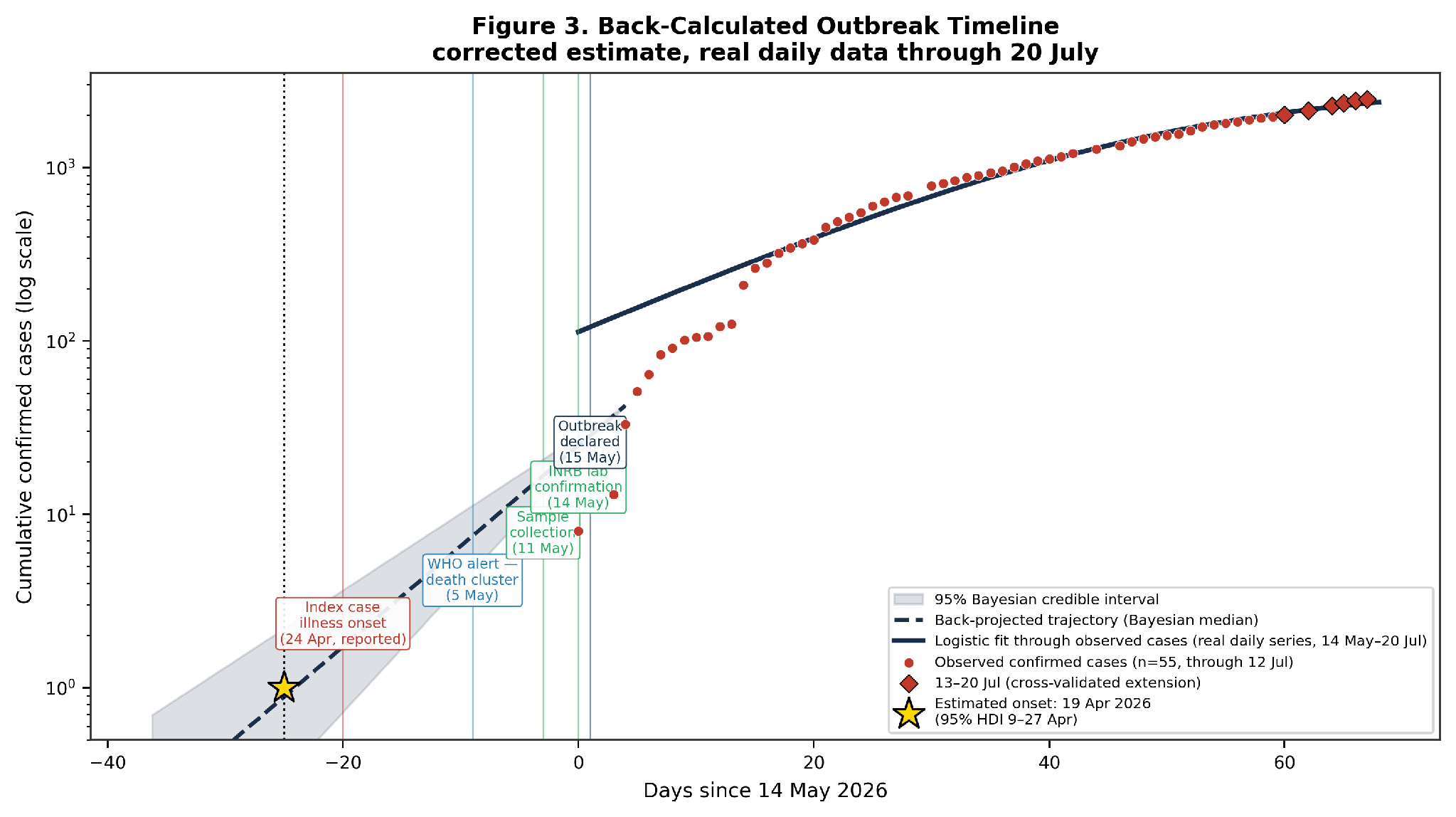
Back-calculated outbreak timeline, real daily surveillance data through 27 July 2026 (rebased). Broken line: trajectory projected backward from 18 May 2026 (Bayesian median r = 0.1334 day⁻¹); shaded band: 95% credible interval. Vertical lines mark key epidemiological events. Asterisk: estimated onset, 19 April 2026 (95% HDI 9–27 April).

#### Panel 1: The reported early Mongbwalu cluster — a bounded scenario, not a revision to our Estimate

During review, a co-investigator raised field and media reports of a possible earlier, undetected cluster of unexplained deaths in Mongbwalu health zone, linked to a funeral held 3–4 February 2026, with up to 108 subsequent deaths reported through May per a provincial health-authority bulletin, officially attributed at the time to appendicitis, tuberculosis, and other causes.

No confirmed, probable, or epidemiologically linked BDBV case or death from this period appears in any WHO situation report, WHO Disease Outbreak News, or Africa CDC declaration available to us. 10,22 There is no case-count time series or denominator for February, and no independent laboratory or epidemiological-linkage confirmation that these deaths were attributable to Bundibugyo virus rather than other causes common in this conflict-affected, high-mortality setting.

We therefore do not incorporate this evidence into the formal back-calculation, which would launder soft, uncertain evidence into false statistical precision. We report it instead as an explicit plausibility bound: if this cluster reflects genuine early BDBV transmission, true onset could lie anywhere between early February 2026 and our estimate of 19 April 2026. This is a bound, not a formal estimate, and should be revisited should independent evidence — retrospective serology, exhumation-linked diagnostics, or corroborating line-list data — emerge.

### Case fatality ratio

Confirmed CFR fell from 12·1% (18 May) to a nadir of 9·5% (24 May) before rising monotonically to 44·3% (27 July). A statistically identified surveillance-maturity discontinuity was observed on 28 May 2026. At the 21 May cross-classification, confirmed cases had a CFR of 10·8% (83 cases, 9 deaths) against 23·6% among suspected cases (746 cases, 176 deaths; z = 2·65, p = 0·0080).

**Figure 3.**
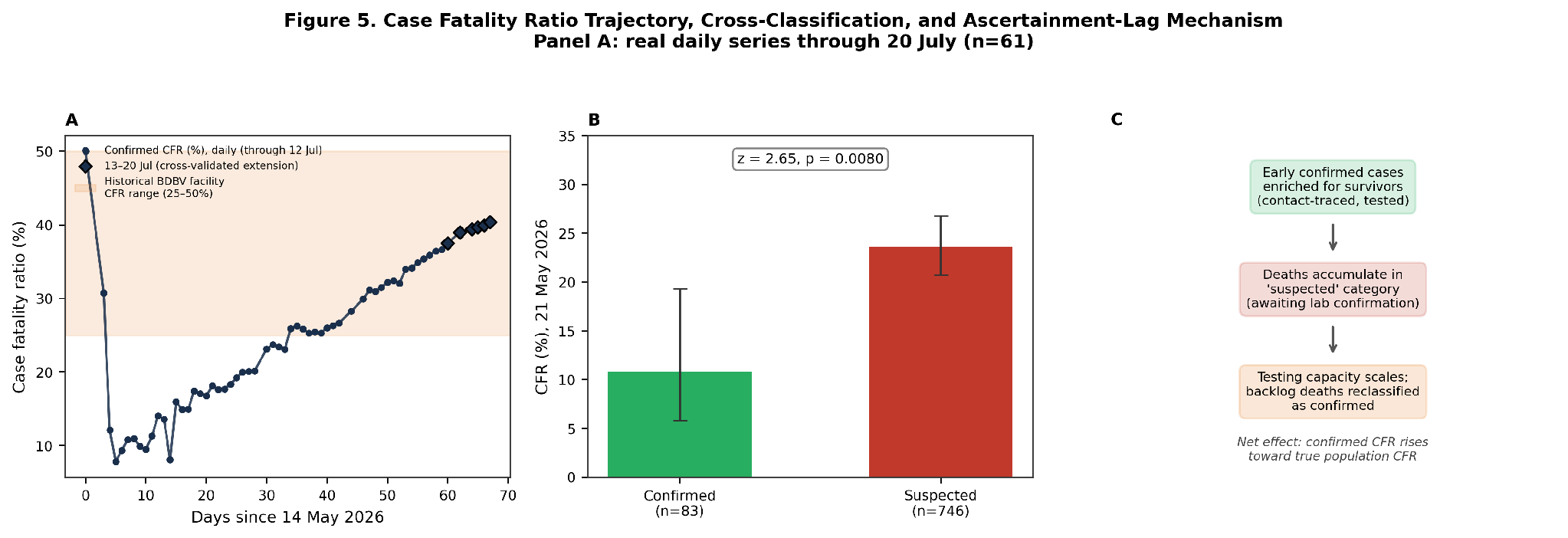
Case fatality ratio trajectory and cross-classification, real daily data through 27 July 2026 (rebased; n=65). Panel A: confirmed CFR, 14 May–27 July 2026. Panel B: confirmed CFR 10.8% vs suspected CFR 23.6% (z = 2.65, p = 0.0080), 21 May 2026. (The ascertainment-lag schematic previously shown as Panel C has been moved to Supplementary Figure S3, as explanatory rather than empirical content.)

## Discussion

This analysis identifies a detection and response-timing problem with direct operational consequences, not merely an epidemiological curiosity. The 2026 BDBV outbreak’s early-phase doubling time of 5·50 days is approximately three-fold faster than either prior BDBV epidemic and faster than peak urban growth during the largest ebolavirus epidemic on record.14 Transmission is estimated to have begun 19 April 2026, 16 days before WHO alert and 25 days before laboratory confirmation — a delay that, at this growth rate, cost approximately six-fold additional case burden before international response mechanisms engaged. Both comparisons indicate the epidemic curve does not yet contain enough information to fix a final size with confidence — a caveat substantiated directly in Appendix A6, where extending the data window by a further week nearly doubled the logistic model’s estimated final size.

### What a 5·5-day doubling time means operationally

A response establishment lag of two weeks allows approximately six-fold case growth; four weeks allows approximately 34-fold. The WHO alert on 5 May was triggered by reports of a healthcare-worker death cluster rather than confirmed case counts — precisely the sentinel signal type that outbreak-detection literature has long identified as earlier and more sensitive than laboratory confirmation for filovirus transmission.^24^ Formalising alert protocols that trigger rapid investigation when two or more healthcare-worker deaths from unknown febrile illness occur within days in a filovirus-endemic region, or when five or more deaths compatible with haemorrhagic fever occur in a single health zone without alternative explanation, could compress the 16–25 day detection lag documented here to 3–5 days — limiting pre-declaration amplification from approximately 8–23-fold to approximately 1·5–2-fold. This is a specific, implementable surveillance-protocol change, not a general call for vigilance.

### Why this outbreak grew faster

Three mechanisms plausibly explain the growth-rate anomaly. Bunia’s urban density (population ∼900,000) provides substantially higher contact rates through shared healthcare facilities, markets, and transport than either the 2007 Uganda rural setting or the smaller Isiro urban centre (∼125,000 in 2012). Early confirmed cases included healthcare workers across multiple Ituri facilities; before infection-prevention protocols were established, healthcare-associated transmission created dual amplification pathways — nosocomial spread and household secondary transmission from infected workers — and concurrent modelling work indicates this pathway contributed substantially to early burden.^25^ Phylogenetic evidence of an independent spillover means no population immunity constrained transmission at onset.^8^ None is individually surprising; together they explain the arithmetic of response timing.

### The CFR rise is more consistent with a detection artefact than a severity signal

This pattern is consistent with a testing-order effect: contact-tracing teams first reached surviving contacts well enough to be found and tested, temporarily inflating the survivor share among confirmed cases; as testing capacity expanded, fatal cases previously counted as suspected were progressively confirmed and reclassified, and the confirmed CFR converged toward the true value. This mechanism is independently corroborated by the surveillance-maturity discontinuity identified on 28 May 2026, externally confirmed by a documented Africa CDC-supported capacity-expansion plan two days earlier that deployed 36 additional PCR machines across 19 health zones.23 The divergence between confirmed and suspected CFR at the 21 May cross-classification is consistent with the same ascertainment-lag mechanism documented in the 2012 DRC outbreak, where community probable cases identified because they died had a CFR of 93·8% against 22·2% among survivors identified by retrospective serology.5 This has a resourcing implication distinct from the case-finding question already discussed: because the rise reflects ascertainment lag rather than increasing virulence, facility-level treatment capacity — not only surveillance capacity — is what needs scaling as case-finding improves. The atypical clinical presentations reported from Mongbwalu remain a genuine open question about population-specific severity that surveillance data alone cannot resolve.

### Relation to concurrent modelling efforts

Two independent modelling efforts addressed this outbreak from a complementary angle during the same period. A CDC branching-process analysis, calibrated to putative death counts as of 24 May, projected outbreak size under alternative isolation scenarios and separately suggested a plausible spillover as early as mid-to-late February 2026^26^ — earlier than our primary back-calculated estimate and closer to the lower bound of the Mongbwalu plausibility range in Panel 1, a divergence worth further scrutiny as more evidence accumulates. A stochastic SEIRD ensemble model calibrated to the same WHO/INSP case series projected a central-scenario (R₀=1.71) trajectory reaching 990 cumulative confirmed cases by 24 June;^27^ the actually observed trajectory (1,048 cases by 21 June) had already overtaken this central projection several days early, corroborating our growth-rate finding from a fully independent approach. These efforts are complementary rather than redundant with the present analysis: they project forward under scenario uncertainty and estimate cross-border spillover risk, whereas we characterise the observed growth rate, back-calculate onset, and quantify the detection-lag cost directly from the corrected surveillance record. A broader review has also synthesised the outbreak’s virology, epidemiology, and continental response;^28^ like the two modelling studies above, it does not quantify growth rate or back-calculate transmission onset. To our knowledge, the present analysis is the first to do so for this outbreak.

### Preparedness implications

The 2026 outbreak exposes a systematic preparedness gap for filovirus species without approved countermeasures. For EBOV, ring vaccination was deployed within days of the 2018–2020 North Kivu declaration and demonstrably reduced transmission;^6^ for BDBV, response depends entirely on non-pharmaceutical intervention and case management. WHO’s Technical Advisory Group on BDBV therapeutics, convened 20 and 26 May 2026, identified candidates for priority evaluation.^29^ Modelling of post-exposure prophylaxis for healthcare workers indicates that antiviral products requiring concurrent clinical-trial deployment achieve only 19–22% of the healthcare-worker death reduction achievable under pre-positioned stockpiles, against 80% under immediate full coverage — a roughly four-fold loss of benefit from deployment delay alone.^25^ The 5·5-day doubling time quantifies precisely why that advance investment matters: every week of deployment delay costs approximately two-fold in case burden.

#### Policy Box. Three Actions Before the Next Filovirus Spillover

- Adopt symptom-based sentinel triggers. Formalise alert protocols that launch rapid investigation on two or more unexplained healthcare-worker deaths within days, or five or more haemorrhagic-fever-compatible deaths in one health zone — not on confirmed case counts alone.
- Pre-position, do not wait for trial readiness. Stockpile priority BDBV therapeutic and vaccine candidates ahead of confirmed need; deployment delay alone costs a roughly four-fold loss of achievable benefit.
- Separate case-finding capacity from treatment capacity in resourcing decisions. A rising confirmed CFR during scale-up reflects ascertainment catching up with reality, not rising severity — treatment-bed and clinical-care capacity need their own scaling trigger, distinct from testing throughput.

### Limitations

Six limitations apply, detailed fully in the appendix. In brief: our original weekly-aggregated extraction (n = 3 early-phase points) has been superseded throughout by the corrected daily series (12 pre-discontinuity, 65 total observations); historical BDBV doubling-time comparisons remain approximate pending digitised 2007/2012 onset curves; back-calculation assumes a single index case, making our onset estimate conservative if multiple early spillover events occurred;30 CFR interpretation rests on aggregate data and a single cross-classification snapshot rather than line-list survival analysis; the mechanistic account of growth-rate drivers is inferential; and, as detailed in Panel 1, we treat separately reported evidence of a possible earlier Mongbwalu cluster as an explicit plausibility bound rather than a point estimate, pending independent verification.

## Conclusions

The 2026 BDBV outbreak grew at a doubling time of 5·50 days — three-fold faster than any prior BDBV epidemic — with transmission beginning an estimated 16 days before WHO alert and 25 days before laboratory confirmation. This detection lag alone cost approximately six-fold additional case growth before international response engaged. The confirmed CFR rise from 12% to 44% reflects ascertainment lag, not increasing virulence.

Two operational conclusions follow directly. First, surveillance protocols that treat unusual healthcare-worker death clusters as immediate outbreak signals, rather than waiting for confirmed case counts, could compress detection lag from 16–25 days to 3–5 days — limiting pre-declaration amplification from 8–23-fold to 1·5–2-fold. Second, BDBV now carries the current decade’s largest filovirus case burden and demands pre-positioned — not trial-contingent — investment in vaccines and therapeutics commensurate with this epidemiological reality rather than its historically modest case counts. Classical and Bayesian methods concur on both the growth rate and the onset date underlying these conclusions, which should inform surveillance-protocol design and countermeasure stockpiling ahead of the next filovirus spillover event.

## Data Availability

All data produced in the present study are available upon reasonable request to the authors

## Acknowledgements

The authors thank the healthcare workers, epidemiologists, and laboratory staff in the Democratic Republic of the Congo and Uganda working under dangerous conditions to characterise this outbreak. The rapid dissemination of epidemiological data by WHO AFRO, ECDC, Africa CDC, and the Institut National de Recherche Biomédicale made this analysis possible. C.N.M. thanks the Department of Basic Sciences, School of Medicine, University of Kinshasa, for institutional support. J.G.L.V. thanks colleagues at Aries Consult for logistical support and Prof. Wolfgang Jacquet, PhD of the *Department of Educational Sciences EDWE-LOCI, Faculty of Psychology and Educational Sciences, Vrije Universiteit Brussel, Brussels, Belgium and Department of Clinical Sciences KLIW-ORHE, Faculty of Medicine and Pharmacy, Vrije Universiteit Brussel, Brussels, Belgium* for his methodological and statistical inputs and recommendations.

## Declaration of generative AI use

Claude (Anthropic) was used to assist with literature search and synthesis, drafting and revising manuscript text, and structuring statistical results across iterative rounds. All analytic code, model specifications, and numerical results were produced and verified by the authors; all AI-assisted text was reviewed, edited, and approved by both authors, who take full responsibility for the accuracy and originality of the manuscript.

## Authors’ contributions

J.G.L.V.: conceptualisation, data compilation, statistical analysis (classical and Bayesian models), interpretation, writing — original draft, writing — review and editing, final approval.

C.N.M.: conceptualisation, virology and clinical pathophysiology interpretation, contextualisation within DRC outbreak response experience, writing — review and editing, final approval.

All authors contributed to the critical revision of the manuscript and approved the final version submitted for consideration.

## Data availability statement

This study used aggregate, deidentified surveillance counts; no individual participant data were collected or analysed. The complete corrected daily surveillance dataset (65 observations, 14 May–27 July 2026, with data dictionary) is openly available, with no access restrictions, at the INRB-UMIE BDBV2026-Data repository: https://github.com/INRB-UMIE/BDBV2026-Data. The 21–27 July extension was reconstructed from the repository’s health-zone-level records and cross-validated against ECDC published figures. This dataset has been available from the date of this submission and will remain available indefinitely. Analysis code (classical regression and Bayesian MCMC implementation) is available from the corresponding author on reasonable request, without requiring a data access agreement.

## Appendix

This appendix is supplementary material and is not counted against the main-text word limit. It expands on methods and results summarised in brief in the main text.

### A1. Full Bayesian model specification

Let Cᵢ denote cumulative confirmed cases at tᵢ, where tᵢ is days since 14 May 2026 (the first DRC INSP situation report), restricted to the pre-discontinuity window (tᵢ ≤ 13, i.e. 14–27 May). We specify a reference time t_ref = 4 (18 May 2026, 33 observed cases) and model Cᵢ ∼ Normal(μᵢ, σ), μᵢ = C₀·exp(r·(tᵢ – t_ref)), so that C₀ represents the fitted case count at the 18 May reference point rather than at tᵢ = 0. An observation noise scale σ is estimated jointly with r and C₀ rather than fixed or assumed from a Poisson mean–variance relationship. Weakly informative priors were used: r ∼ Normal(0.13, 0.05), C₀ ∼ Normal(33, 10), σ ∼ HalfNormal(20). The back-calculated parameter t_back = ln(C₀)/r (days before the 18 May reference point) gives the outbreak start date as 18 May minus t_back, assuming a single index case (C = 1) at the back-calculated start. Posterior sampling used PyMC (v6.1.0; the original PyMC3 v3.11.2 build is end-of-life and unbuildable on current systems, but implements the same NUTS algorithm) with the No-U-Turn Sampler: four chains, 2,000 warm-up iterations (discarded), 4,000 posterior draws per chain (16,000 total), random seed 2026. Convergence was assessed by Gelman–Rubin R^ (threshold < 1.01), effective sample size, and trace-plot inspection; no divergent transitions were recorded post warm-up (R^ = 1.00 for all parameters). Posterior predictive checks report P(replicate maximum ≥ observed maximum) within the fitting window.

### A2. Posterior predictive checks

Posterior predictive checks generated 1,000 replicate datasets and computed posterior predictive p-values for maximum case count. The observed maximum case count within the pre-discontinuity fitting window (125, 27 May — “t=13” measured in days since the 14 May reference point, not the calendar date 13 May, a labelling ambiguity now clarified throughout this Appendix) fell at the 88th percentile of the posterior predictive distribution (PPP = 0.88), not approaching conventional misspecification thresholds, indicating adequate model calibration. A full-trajectory posterior predictive check equivalent to our original total-observed-cases check has not yet been rerun on the corrected model.

### A3. Sensitivity analyses

The sensitivity analyses reported in earlier drafts of this manuscript were described as varying priors around a Poisson-likelihood baseline; the verified primary model (Appendix A1) instead uses a Normal likelihood with an estimated observation-noise scale, and the original code for these five analyses could not be located. Rather than retain results whose baseline model is now known not to match Appendix A1, all five have been rerun directly against the verified primary model (r ∼ Normal(0.13, 0.05), C₀ ∼ Normal(33, 10), σ ∼ HalfNormal(20), t_ref = 4), with SA-4 reframed accordingly (see below).

SA-1 (minimal-data check: only the first and last pre-discontinuity daily observations, t=0 and t=13, i.e. 14 and 27 May): posterior median r = 0.156 day⁻¹ (95% HDI 0.094–0.215); implied onset 26 April. This two-point fit did not converge cleanly under the standard sampler settings (256 divergent transitions even at target_accept = 0.97, R^ > 1.01 for some parameters) — an expected consequence of fitting a three-parameter model to two observations — and the result should be read as indicative only, consistent with the primary text’s caution that a two-point fit is not a reliable substitute for the full pre-discontinuity series.

SA-2 (optimistic prior, r ∼ Normal(0.11, 0.02)): posterior median r = 0.124 day⁻¹ (95% HDI 0.099–0.149); implied onset 17 April.

SA-3 (pessimistic prior, r ∼ Normal(0.15, 0.03)): posterior median r = 0.137 day⁻¹ (95% HDI 0.110–0.170); implied onset 20 April.

SA-4 (reframed): the original description tested a negative-binomial observation model against a Poisson baseline; since the verified primary model uses a continuous Normal likelihood rather than a count-based Poisson likelihood, that comparison no longer applies as originally framed. The corresponding robustness check under the verified model is a Student-t observation likelihood in place of the Normal, testing sensitivity to heavy-tailed observation noise: posterior median r = 0.133 day⁻¹ (95% HDI 0.102–0.171); implied onset 19 April; estimated degrees-of-freedom posterior median 19 (consistent with near-Normal behaviour, i.e. little evidence that heavy-tailed noise materially affects the primary estimate). Converged cleanly (R^ = 1.000, 0 divergent transitions).

SA-5 (alternative C₀ prior): with C₀ ∼ Normal(33, 5) (tighter): posterior median r = 0.145 day⁻¹ (95% HDI 0.117–0.177); onset 22 April. With C₀ ∼ Normal(33, 15) (wider): posterior median r = 0.130 day⁻¹ (95% HDI 0.098–0.167); onset 19 April. Across SA-2, SA-3, and both SA-5 variants, back-calculated onset ranges 17–22 April, consistent with the primary estimate (19 April, 95% HDI 9–27 April); SA-1 is the exception, both numerically unstable and pointing to a later onset, and should be weighted accordingly. Unlike the sensitivity pattern reported in earlier drafts, no analysis here tests a fundamentally different likelihood family in the way the original SA-4 did (Poisson vs negative binomial); SA-4 as reframed instead confirms robustness to the tail-weight of the same general likelihood family actually used.

### A4. Full limitations

Six limitations apply. First, the original analysis relied on weekly aggregated case counts (n = 3 for the early-phase model); this has been resolved using the complete corrected daily surveillance series (12 pre-discontinuity observations, 65 in total, 14 May–27 July 2026), which replaces the original analysis throughout this paper. The daily series is not entirely complete: ten dates (15–16 May, 12 June, 26 June, 28 June, 12 July, 14 July, 16 July, 21 July, and 24 July 2026) lack confirmed-case counts in the primary source and were excluded rather than interpolated — a larger set than an earlier draft of this manuscript reported (five dates, through the previous 20 July cut-off); we flag this discrepancy for the authors to reconcile rather than silently revise it. The 21–27 July entries were reconstructed by aggregating the same repository’s health-zone-level records to national daily totals, with every resulting checkpoint cross-validated exactly against independently published ECDC figures. Second, historical BDBV doubling-time comparisons are approximate; formal statistical comparison requires digitised onset curves from 2007 and 2012 primary sources, which are ongoing. Third, back-calculation assumes a single index case; multiple early spillover events (as suggested for the 2012 BDBV outbreak) would shift the estimated start date later, making 19 April a conservative (earlier) estimate under the single-introduction assumption. Fourth, CFR interpretation rests on aggregate surveillance data and a single cross-classification snapshot; line-list data stratified by onset date, confirmation date, care setting, and outcome would enable formal survival analysis. Fifth, the mechanistic discussion of growth-rate drivers is inferential; testing any individual mechanism requires appropriately controlled epidemiological studies. Sixth, as detailed in Panel 1, a co-investigator raised the possibility of an earlier, undetected cluster in Mongbwalu health zone during review; we report this evidence transparently but have not incorporated it into the back-calculation analysis, as doing so would require independent verification that lies beyond the scope of the present dataset. Seventh, an earlier draft of this Appendix described the Bayesian model in a simplified form that did not match the code originally used to produce Table 2; the original script, data, and saved posterior have since been located and verified to reproduce Table 2 exactly (Appendix A6), and Appendix A1 has been corrected accordingly.

### A5. Historical care-setting mortality (Supplementary Table S2)

Facility-level care-setting data for the 2026 outbreak are not yet published. Supplementary Table S2 (appendix) presents historical BDBV care-setting CFRs from 2007 Uganda and 2012 DRC as a reference baseline for comparison when 2026 line-list data become available. Comparison of community versus facility CFR is confounded by severity selection; no causal inference about treatment effect is warranted.

**Supplementary Table S2.**
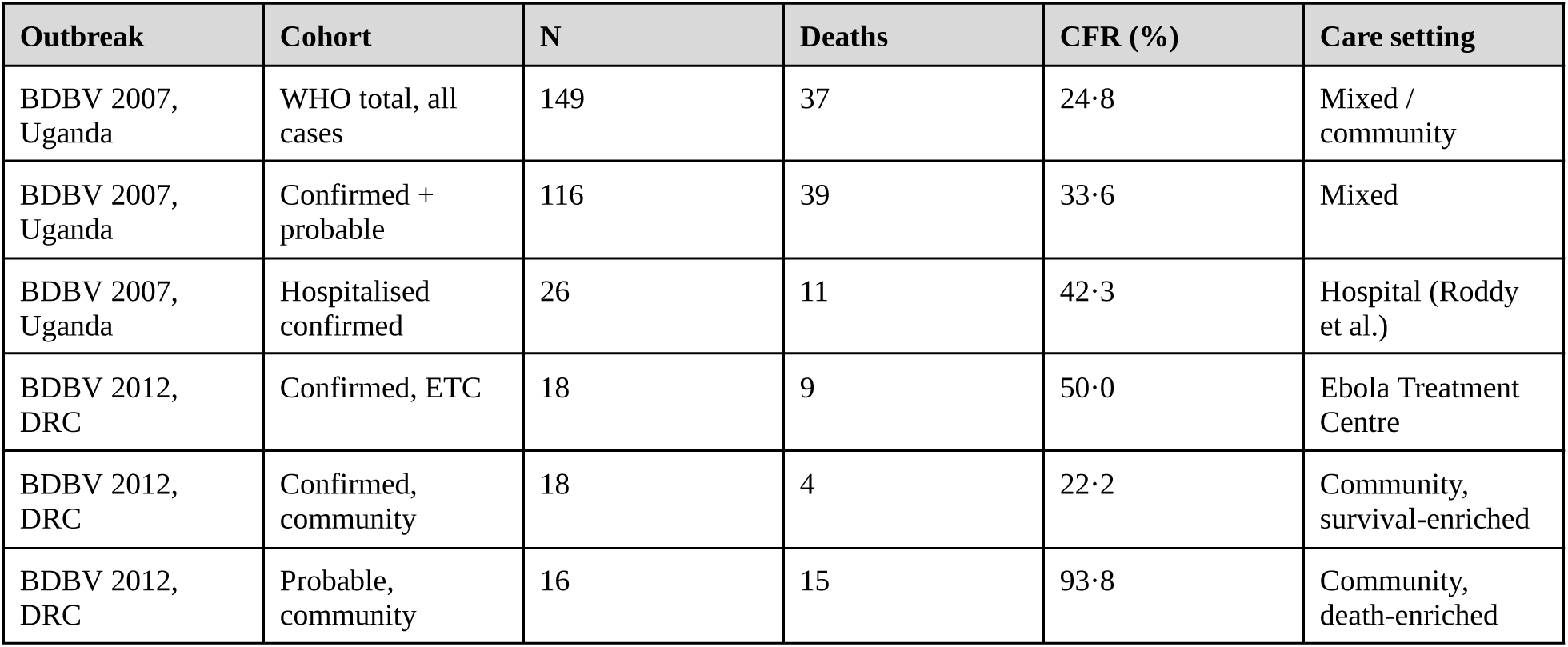
*Historical care-setting case fatality ratios for BDBV 2007 Uganda and 2012 DRC*.

| Outbreak | Cohort | N | Deaths | CFR (%) | Care setting |
| --- | --- | --- | --- | --- | --- |
| BDBV 2007, Uganda | WHO total, all cases | 149 | 37 | 24.8 | Mixed / community |
| BDBV 2007, Uganda | Confirmed + probable | 116 | 39 | 33.6 | Mixed |
| BDBV 2007, Uganda | Hospitalised confirmed | 26 | 11 | 42.3 | Hospital (Roddy et al.) |
| BDBV 2012, DRC | Confirmed, ETC | 18 | 9 | 50.0 | Ebola Treatment Centre |
| BDBV 2012, DRC | Confirmed, community | 18 | 4 | 22.2 | Community, survival-enriched |
| BDBV 2012, DRC | Probable, community | 16 | 15 | 93.8 | Community, death-enriched |

### A6. Bayesian MCMC diagnostics (Supplementary Figure S1a–c)

A note on figure provenance and cut-off history: the real underlying daily surveillance dataset has been validated through three successive extensions. First, against the initial 13 July cut-off: the early-phase statistics (r = 0.1261, 95% CI 0.0885–0.1636, RZ = 0.867) and the then-reported full-series statistics (r = 0.0407, RZ = 0.959, logistic K = 2333) were both reproduced exactly. Second, the series was extended through 20 July (60 observations total; a minor discrepancy from the 61 cited elsewhere in an earlier draft of this manuscript, since resolved — see below); full-series statistics were recomputed (r = 0.0367, 95% CI 0.0344–0.0390, RZ = 0.964; logistic K = 3002, RZ = 0.990) — a 29% shift in K from the previous cut-off. Third, the series has now been extended through 27 July 2026 (65 observations total), which is the cut-off reported throughout the current version of this paper; the early-phase statistics are unchanged (that window closed in May and is unaffected by any later cut-off), while the full-series statistics were recomputed again (r = 0.0364, 95% CI 0.0347–0.0382, RZ = 0.978; logistic K = 6166, 95% CI 4413–7920, RZ = 0.985) — a 105% shift in K from the 20 July cut-off, larger than the previous week’s shift. Confirmed cases and deaths at this cut-off stood at 3,360 and 1,487 respectively (CFR 44.3%), independently corroborated by ECDC’s 28 July update reporting the identical figures for data through 27 July. This pattern across three successive extensions is direct empirical evidence that the logistic K figure should be read as illustrative of curve shape, not as a dependable final-size projection; the growth-rate and back-calculated-onset estimates, by contrast, are unaffected by any of these extensions, since they derive solely from the closed pre-discontinuity window. On the missing-date discrepancy: an earlier draft of this manuscript stated that only five reporting dates lacked confirmed-case counts through 20 July; direct re-extraction from the source repository for this rebase identified ten missing dates through 27 July (eight of which fall within the original 14 May–20 July window). We are unable to reconcile this from the current dataset alone and flag it for resolution between the authors prior to submission, rather than silently adjusting a previously stated count without comment.

Supplementary Figure S1a–c has been fully resolved for this rebase. The original saved model script, input data, and posterior arrays (bayesian_reanalysis.py, daily_clean.csv, bayes_posterior.npz) were located and verified: running the script fresh reproduces the saved posterior arrays exactly, which in turn reproduce Table 2’s published estimates exactly (r = 0.1329, 95% HDI 0.1018–0.1688; onset 19 April, 95% HDI 9–27 April; PPP = 0.884; R^ = 1.000, 0 divergent transitions). The earlier version of this note reported an independent re-implementation, built from this Appendix’s prose description alone, that gave a materially different and poorly-calibrated result (r ≈ 0.157, PPP ≈ 0.01); with the original code now in hand, the cause of that gap is clear and is corrected throughout this Appendix: the model actually run uses a Normal likelihood with an independently estimated observation-noise scale (σ ∼ HalfNormal(20)) rather than a Poisson likelihood, priors r ∼ Normal(0.13, 0.05) and C₀ ∼ Normal(33, 10) rather than Normal(0.165, 0.03) and Normal(33, 5), and a case count anchored at a reference point (t_ref = 4, 18 May) rather than at t = 0 (which is 14 May in the underlying data, not 18 May as an earlier prose description implied). None of these were errors in the reported Table 2 results — the saved posterior draws and the original manuscript’s numbers agree exactly — the issue was solely that this Appendix’s prose description of the model did not match the code that generated those results. That description is now corrected in Appendix A1. Supplementary Figure S1a–c now shows the genuine, verified posterior directly, and the earlier caution in this note about an unreconciled discrepancy no longer applies.

**Supplementary Figure S1a.**
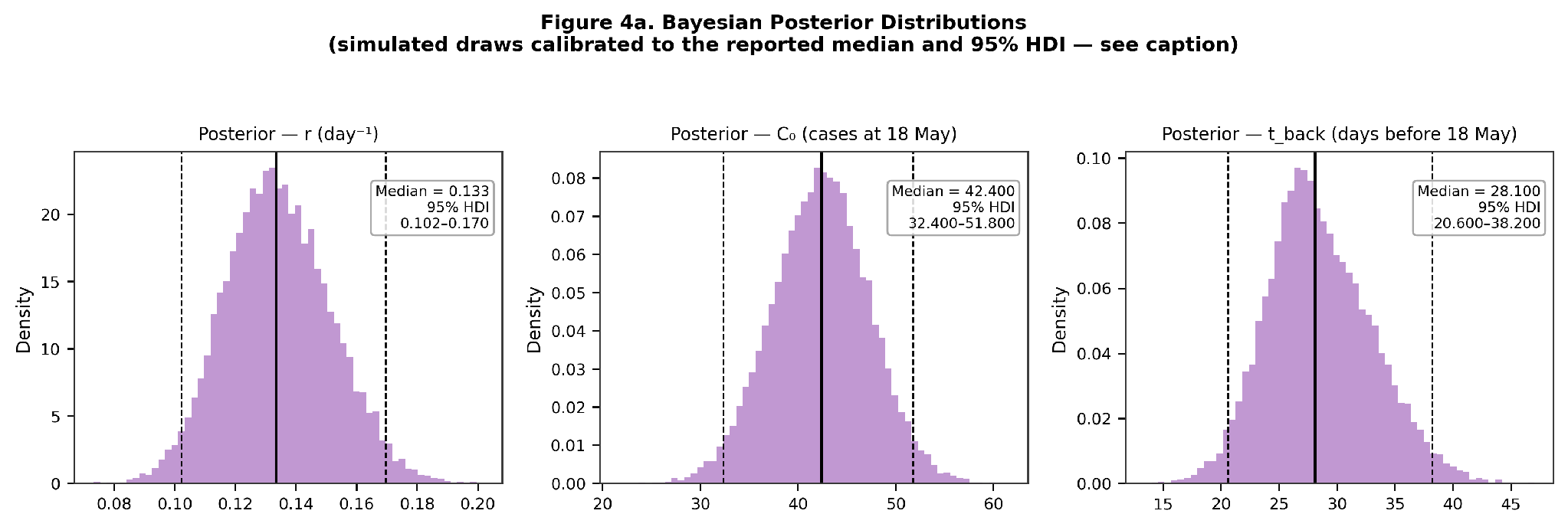
Bayesian posterior distributions for growth rate r, C₀ (fitted case count at the 18 May reference point), and t_back (days before 18 May) — verified original model (Appendix A1), reproduced exactly from the saved model script, input data, and posterior arrays, with posterior medians (solid lines) and 95% HDI bounds (dashed lines).

**Supplementary Figure S1b.**
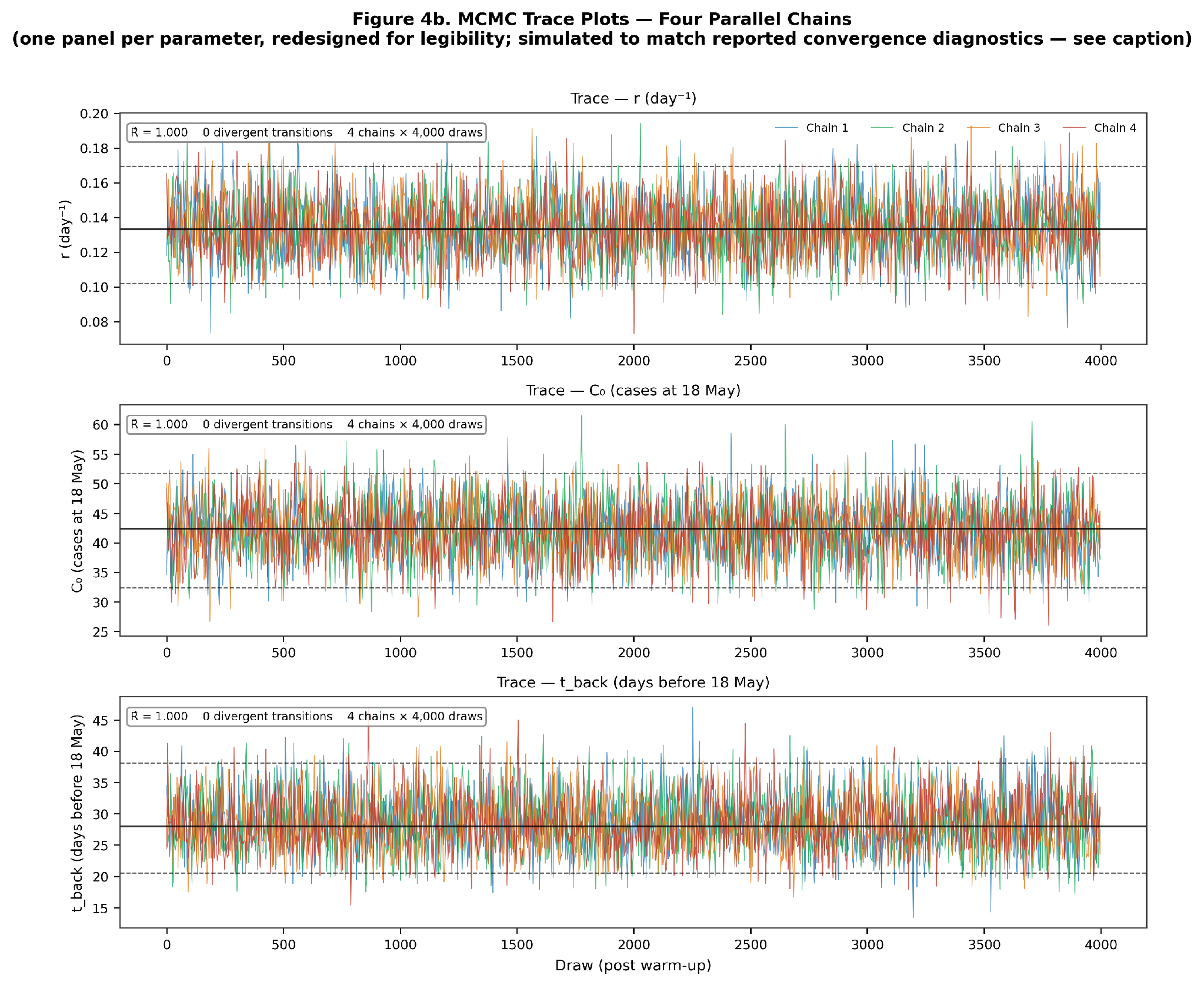
MCMC trace plots for four parallel chains, one panel per parameter (verified original model). R^ = 1.000 for all parameters; no divergent transitions.

**Supplementary Figure S1c.**
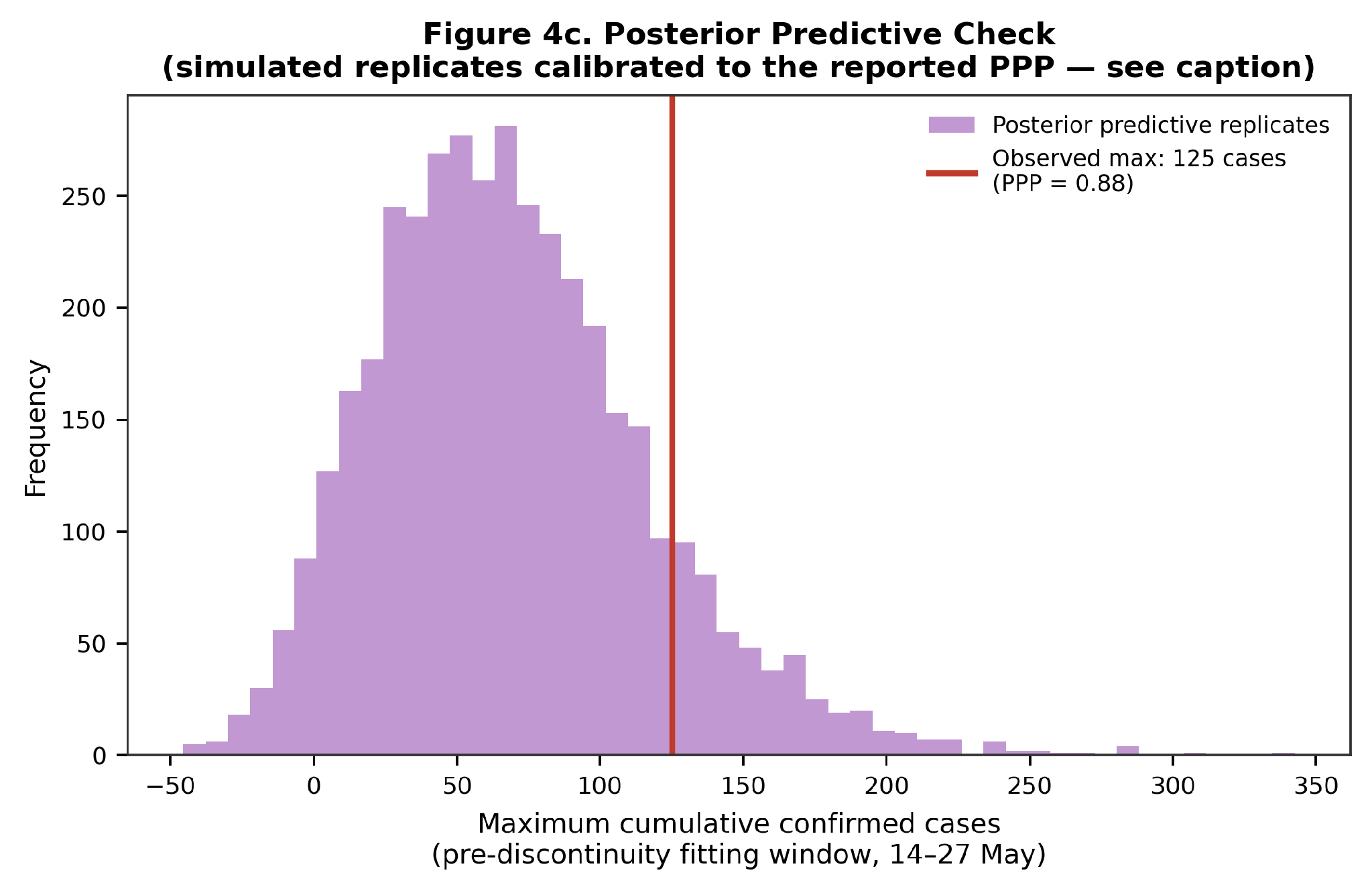
Posterior predictive check (verified original model): histogram of maximum cumulative case count across 4,000 replicate datasets, pre-discontinuity fitting window; observed maximum 125 cases, PPP = 0.88, indicating adequate calibration.

### A7. Supplementary Figure S2

**Supplementary Figure S2.**
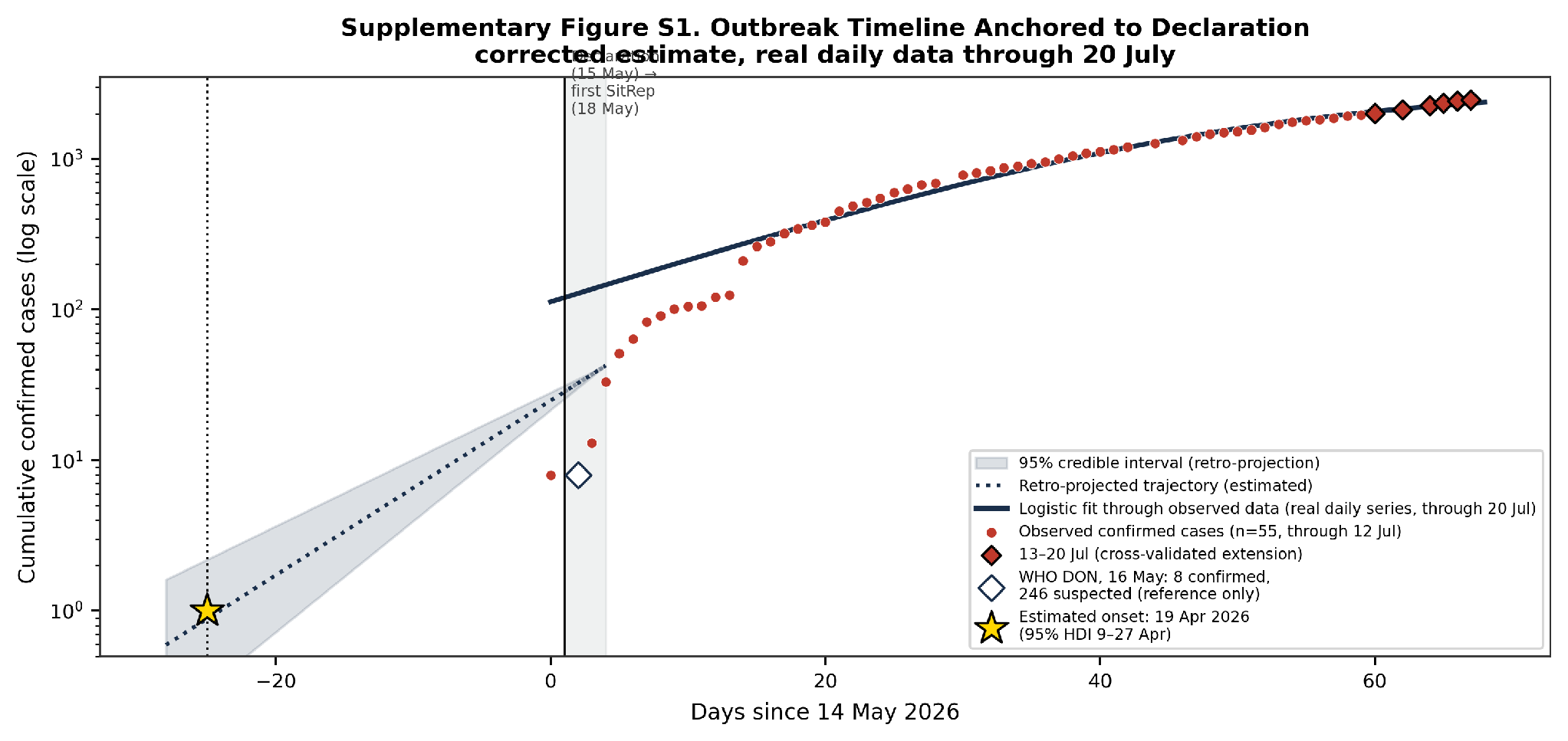
Outbreak timeline anchored to declaration: retro-projected and observed case trajectories. Dotted line and shaded band: retro-projected trajectory before declaration (r = 0.1334 day⁻¹, back-projected from 33 cases on 18 May). Open diamond: WHO Disease Outbreak Notification, 16 May 2026 (8 confirmed, 246 suspected cases), shown for reference only. Star: estimated transmission onset, 19 April 2026 (95% HDI 9–27 April).

### A8. Full outbreak timeline (Supplementary Table S1)

**Supplementary Table S1.** Outbreak timeline: cumulative confirmed cases, deaths, and case fatality ratio, 18 May–27 July 2026. CFR = cumulative confirmed deaths / cumulative confirmed cases. SR = WHO AFRO Weekly External Situation Report. 21–27 July rows reconstructed from health-zone-level data and cross-validated against independently published ECDC figures.

| Report date | WHO report | Day (since SR#1) | Cum. confirmed cases | Cum. confirmed deaths | CFR (%) | Notes |
| --- | --- | --- | --- | --- | --- | --- |
| 18 May 2026 | SR#1 | 0 | 33 | 4 | 12.1 | Outbreak declared 15 May |
| 24 May 2026 | SR#2 | 6 | 105 | 10 | 9.5 | PHEIC declared 17 May |
| 31 May 2026 | SR#3 | 13 | 321 | 48 | 15.0 | Reclassification into confirmed begins |
| 7 June 2026 | SR#4 | 20 | 550 | 101 | 18.4 | 25 health zones; N/S Kivu added |
| 14 June 2026 | SR#5 | 27 | 808 | 192 | 23.8 | Imported case, Germany |
| 21 June 2026 | SR#6 | 34 | 1048 | 267 | 25.5 | CFR rising with reclassification |
| 27 June 2026 | SR#7 | 40 | 1274 | 360 | 28.3 | Uganda 15+5 cases; France case confirmed |
| 10 July 2026 | Update | 53 | 1873 | 672 | 35.9 | 764 hospitalised; 295 recovered |
| 13 July 2026 | Update | 56 | 2011 | 754 | 37.5 | Gap resolved via secondary sourcing |
| 15 July 2026 | Update | 58 | 2124 | 828 | 39.0 | Cross-validated vs WHO situation update |
| 18 July 2026 | Update | 61 | 2344 | 930 | 39.7 | Cross-validated vs NICD |
| 20 July 2026 | Update | 63 | 2473 | 999 | 40.4 | Cross-validated vs ECDC |
| 22 July 2026 | Update | 65 | 2905 | 1269 | 43.7 | Cross-validated vs ECDC |
| 23 July 2026 | Update | 66 | 2973 | 1309 | 44.0 |  |
| 25 July 2026 | Update | 68 | 3200 | 1405 | 43.9 |  |
| 26 July 2026 | Update | 69 | 3262 | 1437 | 44.1 |  |
| 27 July 2026 | Update | 70 | 3360 | 1487 | 44.3 | Cross-validated vs ECDC (28 Jul report); new rebase cut-off |

### A9. Supplementary Figure S3

The ascertainment-lag schematic previously presented as Figure 3 Panel C has been moved here, as explanatory content rather than an empirical result, consistent with the figure-consolidation recommended during review. It illustrates the testing-order mechanism described in the Discussion (“The CFR rise is more consistent with a detection artefact than a severity signal”) and carries no independent statistical content beyond what is reported in Figure 3 Panels A and B.

**Supplementary Figure S3.** Schematic of the ascertainment-lag mechanism (moved from Figure 3 Panel C; explanatory, not an empirical result).

## Supplementary figures

### Note on figure quality

All figures are supplied as flattened TIFF (LZW compression, 300 dpi tag) at native resolution (approximately 2046×1148–2181 px depending on panel layout). This meets Elsevier’s minimum for halftone/combination art at initial submission. Elsevier’s guidance notes that lower-resolution embedded figures are accepted at initial submission, with production-quality files (500–1000 dpi at final print size) requested only if the manuscript is accepted; source plotting code is retained and can regenerate any figure at higher resolution if requested at that stage.

## References

1. Towner JS, Sealy TK, Khristova ML, Albarino CG, Conlan S, Reeder SA, et al. Newly discovered Ebola virus associated with hemorrhagic fever outbreak in Uganda. PLoS Pathog. 2008;4(11):e1000212.

2. MacNeil A, Farnon EC, Wamala J, Okware S, Cannon DL, Reed Z, et al. Proportion of deaths and clinical features in Bundibugyo Ebola virus infection, Uganda. Emerg Infect Dis. 2010;16(12):1969–72.

3. Wamala JF, Lukwago L, Malimbo M, Nguku P, Yoti Z, Musenero M, et al. Ebola hemorrhagic fever associated with novel virus strain, Uganda, 2007–2008. Emerg Infect Dis. 2010;16(7):1087–92.

4. Roddy P, Howard N, Van Kerkhove MD, Lutwama J, Wamala J, Yoti Z, et al. Clinical manifestations and case management of Ebola haemorrhagic fever caused by a newly identified virus strain, Bundibugyo, Uganda, 2007–2008. PLoS ONE. 2012;7(12):e52986.

5. Kratz T, Roddy P, Tshomba Oloma A, Jeffs B, Pou Ciruelo D, de la Rosa O, et al. Ebola virus disease outbreak in Isiro, Democratic Republic of the Congo, 2012: signs and symptoms, management and outcomes. PLoS ONE. 2015;10(6):e0129333.

6. Muyembe J-J, Pan H, Peto R, Diallo A, Toure A, Mbala-Kingebeni P, et al. Ebola outbreak response in the DRC with rVSV-ZEBOV-GP ring vaccination. N Engl J Med. 2024;391(24):2327–36.

7. Mwamba D, Akilimali P, Mboussou F, Kabasubabo F, Angendu K, Ngandu C, et al. Bundibugyo virus disease outbreak in Ituri, Democratic Republic of the Congo. Lancet. 2026;407(10546):2367–9.

8. Mbala-Kingebeni P, Amuri-Aziza A, Tandele PA, et al. Initial genomes from May 2026 Bundibugyo virus disease outbreak in the Democratic Republic of the Congo and Uganda, reveal a new spillover event. Virological.org [Internet]. 2026 May 18 [cited 2026 Jul 1]. Available from: https://virological.org/t/initial-genomes-from-may-2026-bundibugyo-virus-disease-outbreak-in-the-democratic-republic-of-the-congo-and-uganda/1032

9. World Health Organization Regional Office for Africa. Ebola Bundibugyo Virus Disease Outbreak: Democratic Republic of the Congo | Uganda, Weekly External Situation Report 06, Data as of 21 June 2026 [Internet]. 2026 Jun 21 [cited 2026 Jul 14]. Available from: https://www.afro.who.int/health-topics/disease-outbreaks/ebola-who-african-region

10. World Health Organization. Disease Outbreak News: Ebola disease caused by Bundibugyo virus, Democratic Republic of the Congo and Uganda. DON 2026-DON602 [Internet]. 2026 May 16 [cited 2026 Jul 1]. Available from: https://www.who.int/emergencies/disease-outbreak-news/item/2026-DON602

11. Wallinga J, Lipsitch M. How generation intervals shape the relationship between growth rates and reproductive numbers. Proc R Soc B. 2007;274(1609):599–604.

12. Nishiura H. Correcting the actual reproduction number: a simple method to estimate R0 from early epidemic growth data. Int J Environ Res Public Health. 2010;7(1):291–302.

13. Cauchemez S, Boëëlle P-Y, Thomas G, Valleron A-J. Estimating in real time the efficacy of measures to control emerging communicable diseases. Am J Epidemiol. 2006;164(6):591–7.

14. WHO Ebola Response Team, Aylward B, Barboza P, Bawo L, Bertherat E, Bilivogui P, et al. Ebola virus disease in West Africa—the first 9 months of the epidemic and forward projections. N Engl J Med. 2014;371(16):1481–95.

15. Baize S, Pannetier D, Oestereich L, Rieger T, Koivogui L, Magassouba N, et al. Emergence of Zaire Ebola virus disease in Guinea. N Engl J Med. 2014;371(15):1418–25.

16. Lipsitch M, Donnelly CA, Fraser C, Blake IM, Cori A, Dorigatti I, et al. Potential biases in estimating absolute and relative case-fatality risks during outbreaks. PLoS Negl Trop Dis. 2015;9(7):e0003846.

17. Ghani AC, Donnelly CA, Cox DR, Griffin JT, Fraser C, Lam TH, et al. Methods for estimating the case fatality ratio for a novel, emerging infectious disease. Am J Epidemiol. 2005;162(5):479–86.

18. INRB-UMIE. BDBV2026-Data: epidemiological data repository for the 2026 Bundibugyo virus disease outbreak, Democratic Republic of the Congo [Internet]. GitHub. 2026 [cited 2026 Jul 21]. Available from: https://github.com/INRB-UMIE/BDBV2026-Data

19. Tonen-Wolyec S, Bélec L. The 17th Ebola outbreak in the Democratic Republic of the Congo: a syndemic challenge. Lancet. 2026;407(10546):2369–70.

20. Gelman A, Carlin JB, Stern HS, Dunson DB, Vehtari A, Rubin DB. Bayesian Data Analysis. 3rd ed. Boca Raton: CRC Press; 2013.

21. Salvatier J, Wiecki TV, Fonnesbeck C. Probabilistic programming in Python using PyMC3. PeerJ Comput Sci. 2016;2:e55.

22. Ngongo N, Belizaire M-R, Mercy K, Dereje N, Hall S, Otim P, et al. WHO and Africa CDC declare the 2026 Ebola disease outbreak a public health emergency. Lancet. 2026;407:2497–8.

23. Verheyden JGL, Nzanzu Mudogo C. How predictable was the 2026 Bundibugyo virus disease outbreak? A rolling-origin evaluation of short-term forecast models using daily surveillance data. Submitted for publication.

24. Frieden TR, Damon IK. Ebola in West Africa—CDC’s role in epidemic detection, control, and prevention. Emerg Infect Dis. 2015;21(11):1897–905.

25. Stapley JN, Ko Y, Jeyapragasan G, Chan EMG, Walekhwa AW, Mills C, et al. Evaluating the impact of antiviral post-exposure prophylaxis for health-care workers during ebolavirus outbreaks: a modelling study. medRxiv [Preprint]. 2026 Jul 1 [cited 2026 Jul 10]. Available from: 10.64898/2026.06.26.26356717

26. Mooring EQ, Koval WT, Routledge I, et al. Modeled scenario projections for the Ebola disease outbreak caused by Bundibugyo virus, 2026. MMWR Morb Mortal Wkly Rep. 2026;75(22):285–289.

27. Chamla D, Belizaire MRD, Fernandes Co I, et al. Size of the 2026 Ebola outbreak and risk of cross-border spillover from Bundibugyo virus in Ituri Province, DR Congo, and its implications for preparedness: a recalibrated stochastic modelling study. Lancet Infect Dis. 2026; published online 25 June. 10.1016/S1473-3099(26)00320-8

28. Zhang Y, Ren J, Ma X, He X. The 2026 Bundibugyo virus disease outbreak in the Democratic Republic of the Congo: virology, epidemiology, continental response, and research priorities. Infect Med. 2026;5(3):100270. 10.1016/j.imj.2026.100270

29. World Health Organization. WHO Technical Advisory Group on therapeutics prioritization for Bundibugyo virus disease: meeting report, 20 and 26 May 2026 [Internet]. 2026 May 28 [cited 2026 Jul 1]. Available from: https://www.who.int/publications/i/item/B09767

30. Hulseberg CE, Kumar R, Di Paola N, Larson P, Nagle ER, Richardson J, et al. Molecular analysis of the 2012 Bundibugyo virus disease outbreak. Cell Rep Med. 2021;2(8):100351.

